# Evaluation of minimum-to-severe global and macrovesicular steatosis in human liver specimens: a portable ambient light-compatible spectroscopic probe

**DOI:** 10.1101/2023.12.04.23299259

**Authors:** Hao Guo, Ashley E. Stueck, Jason B. Doppenberg, Yun Suk Chae, Alexey B. Tikhomirov, Haishan Zeng, Marten A. Engelse, Boris L. Gala-Lopez, Anita Mahadevan-Jansen, Ian P.J. Alwayn, Andrea K. Locke, Kevin C. Hewitt

## Abstract

**Background & Aims:** Hepatic steatosis (HS), particularly macrovesicular steatosis (MaS), influences transplant outcomes. Accurate assessment of MaS is crucial for graft selection. While traditional assessment methods have limitations, non-invasive spectroscopic techniques like Raman and reflectance spectroscopy offer promise. This study aimed to evaluate the efficacy of a portable ambient light-compatible spectroscopic system in assessing global HS and MaS in human liver specimens.

**Methods:** A two-stage approach was employed on thawed snap-frozen human liver specimens under ambient room light: biochemical validation involving a comparison of fat content from Raman and reflectance intensities with triglyceride (TG) quantifications and histopathological validation, contrasting Raman-derived fat content with evaluations by an expert pathologist and an artificial intelligence (AI) algorithm. Raman and reflectance intensities were combined to discern significant (≥10%) discrepancies in global HS and MaS.

**Results:** The initial set of 16 specimens showed a positive correlation between Raman and reflectance-derived fat content and TG quantifications. The Raman system effectively differentiated minimum-to-severe global and macrovesicular steatosis in the subsequent 66 specimens. A dual-variable prediction algorithm, was developed, effectively classifying significant discrepancies (>10%) between AI-estimated global HS and pathologist-estimated MaS.

**Conclusion:** Our study established the viability and reliability of a portable spectroscopic system for non-invasive HS and MaS assessment in human liver specimens. The compatibility with ambient light conditions and the ability to address limitations of previous methods marks a significant advancement in this field. By offering promising differentiation between global HS and MaS, our system introduces an innovative approach to real-time and quantitative donor HS assessments. The proposed method holds promise of refining donor liver assessment during liver recovery and ultimately elevating transplantation outcomes.

**Lay Summary:** This research explored a portable ambient light-compatible spectroscopic probe to non-invasively assess global and macrovesicular steatosis in human liver specimens. Our findings suggest that this method can be a reliable tool to aid surgeons’ decision-making on a liver’s suitability for transplantation.

**Graphic Abstract:** 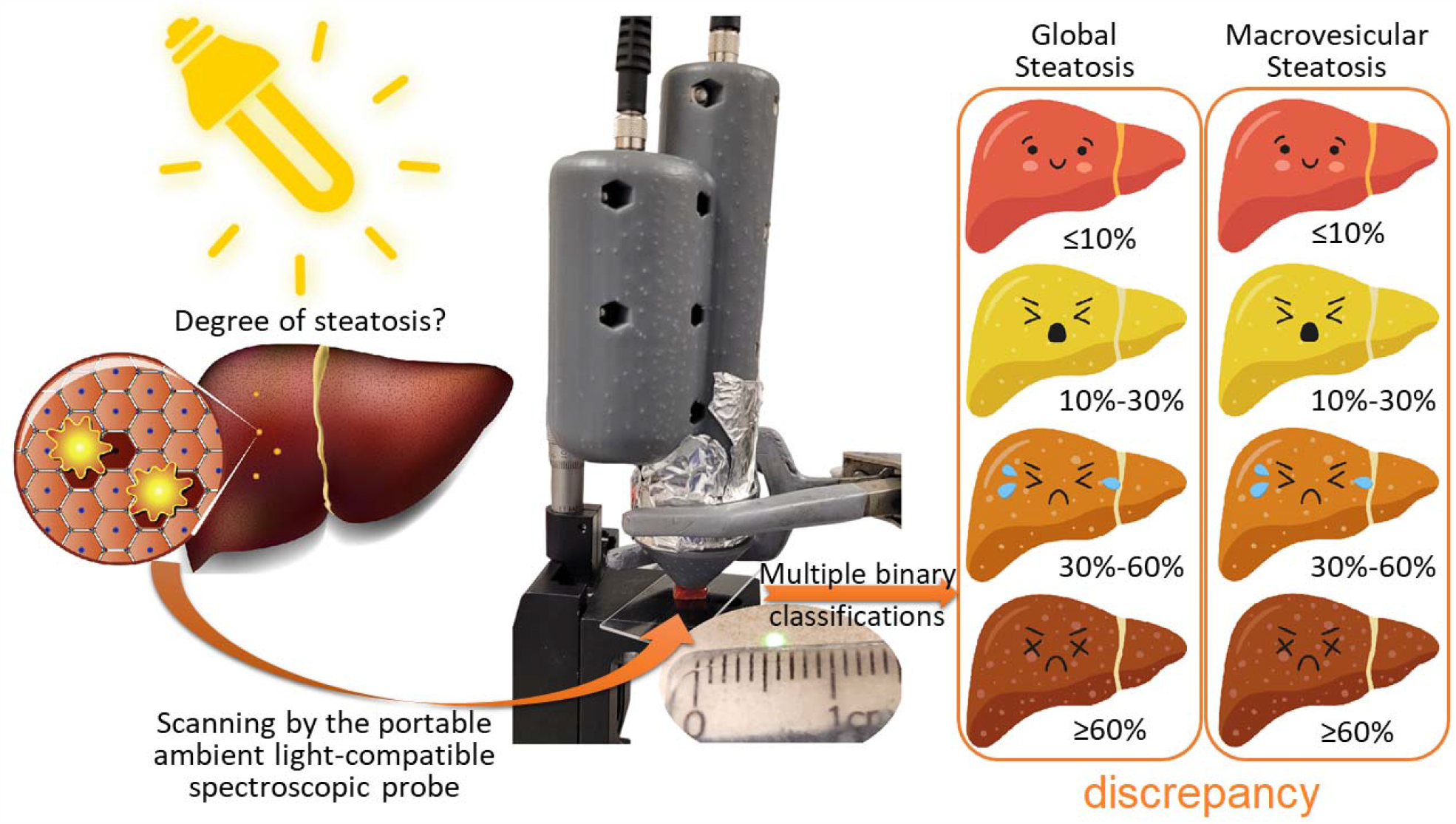

**Highlights:**

- We introduced a portable ambient light-compatible spectroscopic probe which could non-invasively analyze Raman scattering and reflectance of human liver specimens.
- Both biochemical and histopathological approaches were applied to validate the spectroscopic probe.
- Histopathological evaluation covered minimum-to-severe global and macrovesicular steatosis, surpassing previous spectroscopic studies.
- A dual-variable prediction for precise differentiation between global and macrovesicular steatosis offers the potential for enhanced, real-time, and quantitative liver assessments in clinical settings.

## 1 Introduction

Hepatic steatosis (HS), particularly macrovesicular steatosis (MaS) in donor livers, has been found to be linked to a higher likelihood of graft dysfunction following liver transplantation (LT). ^1,2^ Severe MaS (≥60%) is typically viewed as high risk and leads to organs being discarded.^3^ As the donor liver pool expands to accommodate the growing number of patients on waiting lists, it is increasingly essential to conduct thorough assessments of survival benefits among candidates.^4,5^ High-quality estimations of risk factors are crucial, as donors with less favourable characteristics are now more frequently considered for transplantation. The recovery rate of donor livers with moderate (30%-60%) MaS has increased over the years, while other risk factors of mortality or graft loss were mitigated^6,7^. However, the discard rate of moderately and severely steatotic livers is still high because of their higher risk of graft dysfunction.^8,9^ According to a recent study on 17,801 liver transplant recipients, compared with recipients of grafts with 0-10% MaS, the hazard of graft failure was found to be 53% and 25% higher among the recipients of grafts with >30% MaS and 10%-30% MaS, respectively.^10^ Therefore, precise classifications and quantification of steatotic donor livers, alongside a clear-cut threshold at different degrees of MaS, are crucial for optimizing liver allocation and maximizing the survival benefit of liver recipients.

At present, donor livers are assessed macroscopically at the time of recovery.^11^ This assessment is often subjective and dependent on the experience of the donor surgeon. If necessary, frozen sections can be obtained, but this often is associated with significant delays and requires the availability of an expert pathologist.^12^ A tool that can provide objective, quantitative and rapid analysis of (donor) liver fat content is needed.

Optical spectroscopic tools, including infrared (IR), reflectance, and Raman, have shown significant potential in evaluating HS due to their minimally invasive nature and rapid analysis capabilities.^11,13^ Recent studies on both animal models and human livers have demonstrated that IR spectroscopy^14–16^ and reflectance spectroscopy^17,18^ could evaluate various degrees of global HS (with out differentiating MaS); however, those applications required either switching off (directing away) surgical lights or using an invasive optical needle in the operating room (OR). Furthermore, complex spectral analyses of acquired data were a necessity.

Raman spectroscopy offers some distinct advantages. It is sensitive to rotations and vibrations of chemical bonds, making its inherent high molecular specificity ideal for characterizing biological materials.^11,19^ Pre-clinical studies on *ex vivo* rodent liver specimens revealed that Raman spectroscopy could quantify HS in animal models using signals in the high wavenumber region 2800–3000 cm^−1^, and the results agreed well with pathology ratings.^20–22^ Confocal Raman microscopy has been reported to effectively analyze the sizes of lipid droplets in rodent livers.^23–25^ This offers insights relevant to microvesicular steatosis (MiS) and MaS. However, obtaining a Raman image can be considered too time-consuming to be helpful when assessing the HS of donor livers, as each pixel requires spectral analyses.

The exploration of HS in LT using conventional Raman spectroscopy lags IR and reflectance spectroscopy. Hewitt et al. (2015)^20^ and Pacia et al. (2018)^21^ proved the concept that conventional Raman spectrometers could assess the fat content of rodent livers. However, neither of the two studies discussed HS assessment at morphological levels.

Navigating the challenges associated with weak signal detection and fluorescence interferences, we engineered a filter-based 1064-nm Raman system validated through examinations using phantom models and duck liver samples.^26^ In our previous study, employing readily interpretable voltage intensities to quickly quantify the relative fat content within the examined liver samples, this multi-channel system functioned reliably (r^2^ = 0.934) under normal and intense ambient light conditions^26^.

In the present study, we analyzed 95 specimens in two sets from two medical centers, utilizing our filter-based 1064-nm Raman system. For the initial set of 16 specimens, fat contents converted from Raman and reflectance intensities were contrasted with triglyceride (TG) quantification results.

Subsequently, for the more extensive set of 66 specimens, fat contents derived from Raman and reflectance intensities were compared to evaluations of MaS and global HS (including MiS and MaS) performed by an expert pathologist and to determinations of positive pixels made by an artificial intelligence (AI) algorithm.

## 2 Materials and Methods

### 2.1. Optical filter-based Raman system

We used a lab-made optical filter-based Raman system, as previously described^26^. The Raman system integrates a continuous-wave 1064 nm laser to illuminate the samples, a handheld probe and an optical filter system for selecting specific wavelength bands, and a conditioner which employs a phase-sensitive technique detection to amplify and isolate the component of the signal at a specific reference frequency. Laser beam is modulated at a 5600 Hz frequency. The light reflected from the liver specimen is collected, guided back into the system, split into two distinct wavelength ranges (reflectance and Raman) and measured by detectors for further analysis.

We calibrated the Raman system using MRI-calibrated duck fat-agar phantoms with the fat content of 0%-70% before each series of measurements (an example calibration curve is shown in **Figure S1**). The Raman and reflectance measurements followed the methods outlined in Ref. [26] in ambient room light^26^. The Raman and reflectance light intensities were converted to voltage changes by InGaAs photodiodes and a transimpedance amplifier for direct readout.

### 2.2. Design of the study

The study employed a two-stage approach to ascertain the validity of Raman spectroscopy technique in assessing the degree of steatosis in human liver specimens, as demonstrated in **Figure 1**.

**Figure 1.**
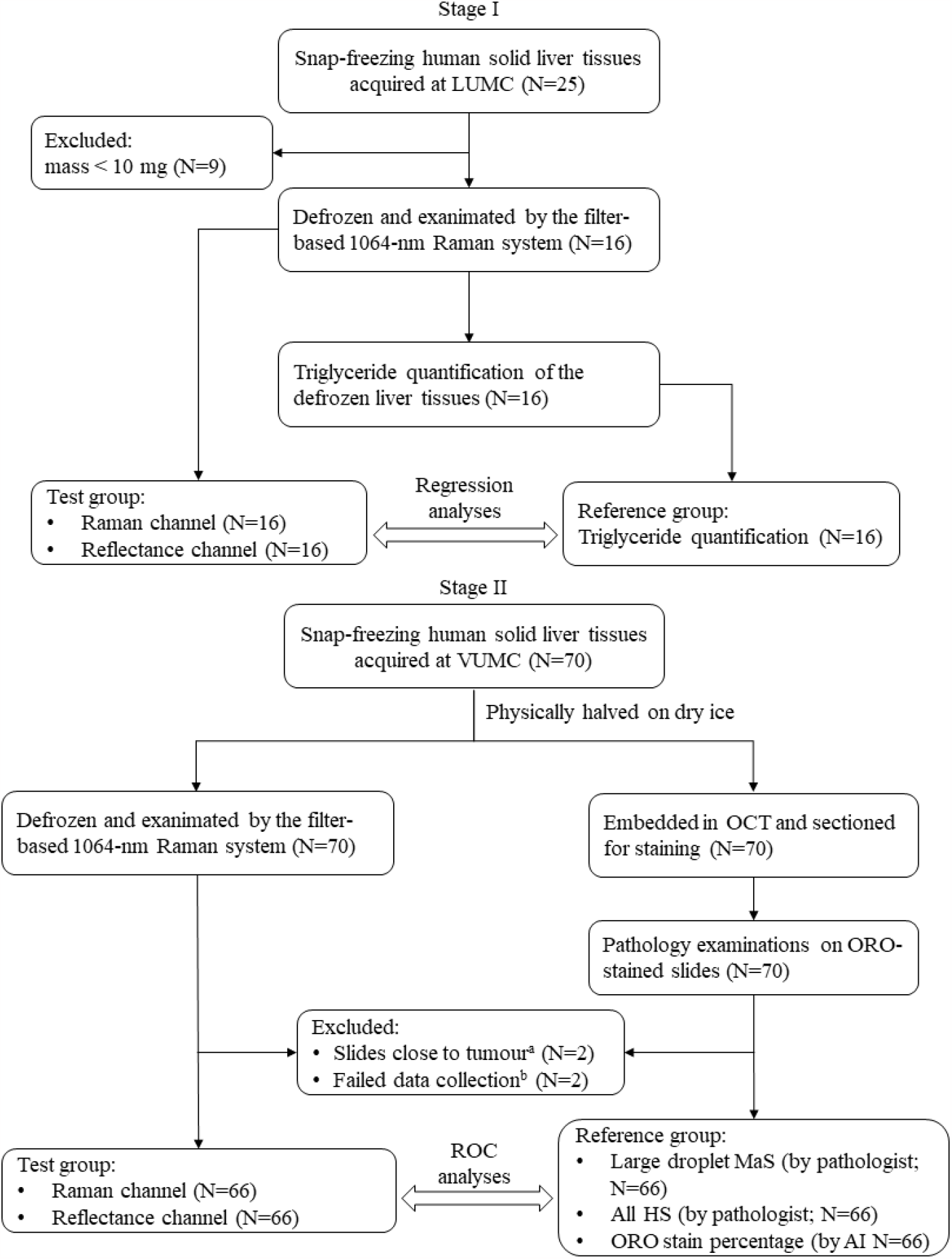
Flow chart of the present study. (a) The specimens had tumours, and the stained frozen section was physically close to the tumour. (b) The SD of voltage signals in either channel was greater than 80 μV.

#### 2.2.1 Stage I: biochemical validation

In the first stage, we studied 25 snap-frozen human liver specimens (weighing between 1.3 mg and 58.1 mg): 24 from remnants of pre- and post-LT wedge biopsies and one from a discarded donor liver at Leiden University Medical Center (LUMC) in August 2022. The specimens were thawed at room temperature and scanned using the Raman system under ambient room light. The liver specimens were refrozen at -80 degrees Celsius and thawed again for TG quantification. Given that the penetration depth of the incident laser beam was at least 1mm,^26^ only specimens heavier than 10 mg (thicker than 1 mm) were included to avoid the insufficiency to quantify TG of low-mass specimens^27,28^. The fat content derived from the analyses using the Raman and reflectance channels was then compared with the TG quantification results to assess the Raman system’s accuracy in steatosis estimation.

#### 2.2.2 Stage II: histopathological validation

In the second stage, at Vanderbilt University Medical Center (VUMC) in November 2022, 70 snap-frozen human liver specimens weighing between 0.24 g and 2.58 g were obtained through the Cooperative Human Tissue Network. These liver specimens were split on dry ice: one half was thawed in cold water and scanned under ambient light using the investigated Raman system, while the other half was cryo-sliced and stained with Oil Red O (ORO) for histological analysis. The Raman system user (HG) and the expert pathologist (AES) performed the Raman and histopathological assessments independently, blinded to the results of each other assessment. Four liver specimens were excluded due to proximity to tumours or due to Raman data collection issues. The Raman results were compared to expert pathologist evaluations of steatosis.

### 2.3. Spectroscopical evaluation of fat content

In the biochemical validation stage: Stage I (Figure **2A**), data collection lasted 10 seconds at 28 recordings per second. Data collection in the histopathological validation stage: Stage II (Figure **2B**) lasted 20 seconds at ten recordings per second.

**Figure 2.**
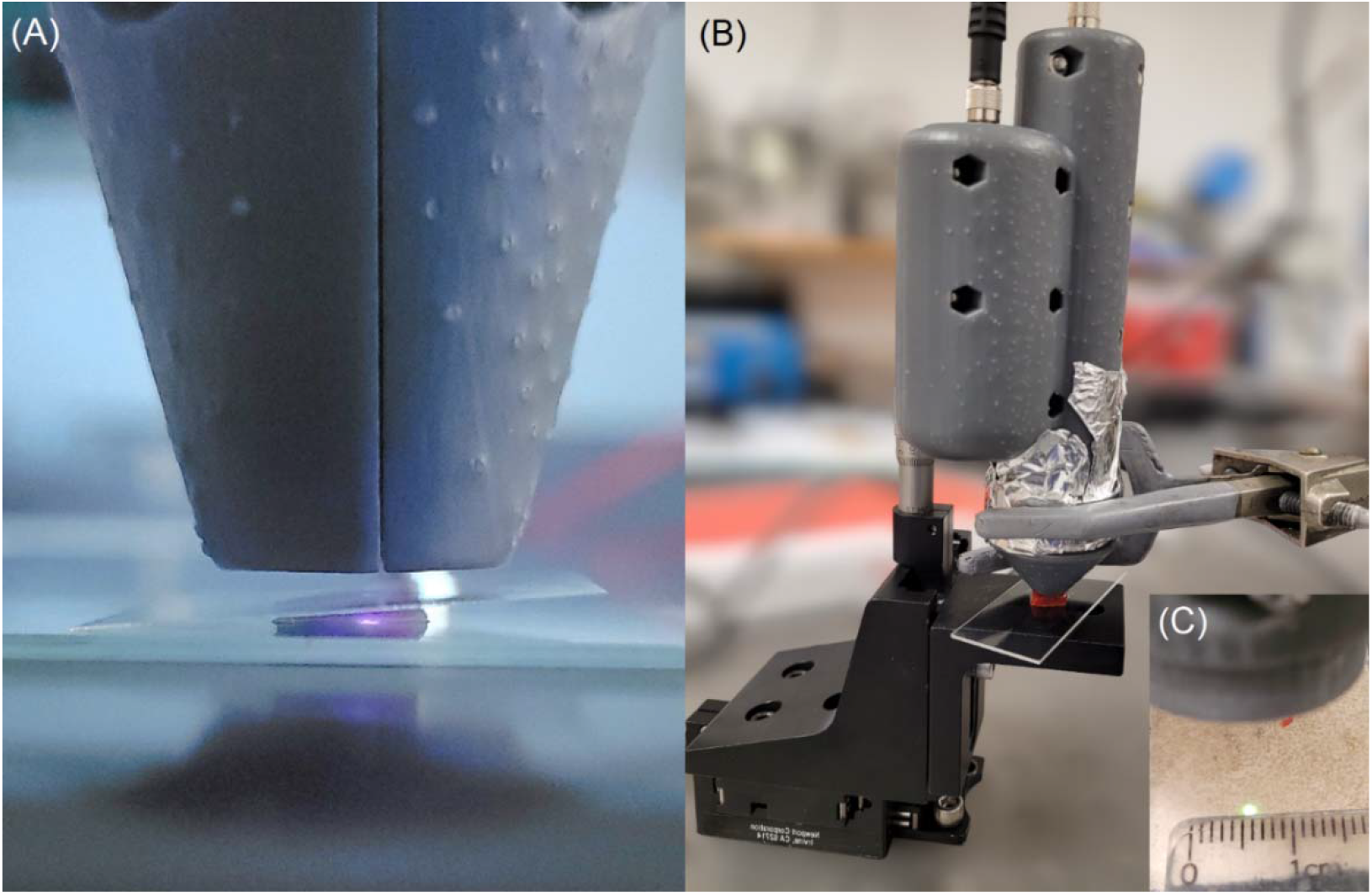
System setup for measurements at (A) the biochemical validation stage and (B) the histopathological validation stage. (An aluminum tape in the picture fixed the reusable tip, so its relative position to the probe did not change.) (C). Laser spot size measurement using a ruler on an infrared viewer card. The distances from the liver specimens and the card to the front lens inside the probe were consistent.

During an entire series of measurements, the handheld probe was fixed on a stand to ensure a consistent distance (which is the front focal length of the front lens) from the liver specimen to the front lens inside the probe. The laser light was directed towards a liver specimen though the handheld probe optics using a multi-modal optical fibre and providing 1.5 mm and 150 mW on sample beam diameter and power, respectively. (Figure **2C**).

### 2.4. Triglyceride quantification

The TG quantification method used in the biochemical validation of this study was based on the liver lipid quantification methods introduced by Nahon et al. (2018)^27^ and de Jong et al. (2022)^28^. TG was extracted from liver specimens higher than 10mg. The liver specimens were homogenized using a potter machine in a solution of 200-500 μL Nonidet™ P 40 Substitute (Sigma-Aldrich Corp., St. Louis, MO, USA), depending on the weight of each specimen. From the homogenate, 10 μL aliquots were retained for protein analyses, and the remaining homogenate was subjected to three cycles of heating in the presence of Nonidet™ P 40 Substitute until the solution became clear, followed by cooling on ice to ensure the complete solubilization of TGs. Any insoluble material was subsequently removed by centrifugation at 13,148 rcf for two minutes. The supernatant, enriched with solubilized TGs, was used for quantification. TG concentrations were determined using the same enzymatic colorimetric assay described by Out et al.^29^, with the absorbance measured at 490 nm and an incubation period of 30 minutes at 37°C. Standard TG samples were employed as a reference during the colorimetric assays.

Protein concentrations in the liver were determined using an assay similar to the Pierce™ BCA Protein Assay Kit (ThermoFisher Diagnostics, Waltham, MA, USA). The TG contents were normalized per unit protein mass by dividing the measured TG concentrations by the corresponding protein concentrations.

### 2.5. Pathological evaluation of steatosis

5-um thick sections were cut from the snap-frozen human liver specimens and placed on microscope slides. The sections were stained with ORO.^30^ ORO specifically stains TGs and cholesteryl oleate in the cytoplasm.^31^ ORO-staining operates well on frozen tissues, and it is commonly used for accurately identifying MiS and MaS.^22,32–34^ MaS typically appears as a single large lipid droplet in a liver cell, pushing the nucleus to the side and being at least as large as the nucleus. On the other hand, MiS was characterized by one or more small lipid droplets inside the cell’s cytoplasm that didn’t shift the nucleus and were smaller than it.

Slides were digitized at 40X magnification in brightfield using an Aperio AT2 scanner (Leica Biosystems, Deer Park, IL, USA). In addition to the pathologist’s evaluation, an AI algorithm assessed the global HS by quantifying the ORO stain percentage based on the stained slide pixel area. As the AI algorithm mechanically summarizes the area of ORO-stained pixels, it cannot quantify MaS based on the Banff consensus of what constitutes a large lipid droplet.^12^

### 2.6. Statistical methods

Stata version 18.0-Basic Edition (StataCorp LLC, TX, USA) was used for statistical analyses. Logistic regression was performed, and performance of the investigated Raman system in HS prediction was evaluated through serial binary classifications and corresponding Receiver Operating Characteristic (ROC) analyses. As a non-parametric test, the Mann-Whitney U test was a used to compare whether there is a significant difference in the distributions of two groups of samples after classification.

## 3 Results

This study included 16 and 66 human liver specimens in the biochemical and histopathological validation stages for analyses. The characteristics of the population are shown in **Table 1**. The ethnicity information was not available in the biochemical validation stage, as it is not routinely captured in the LUMC database.

**Table 1.**
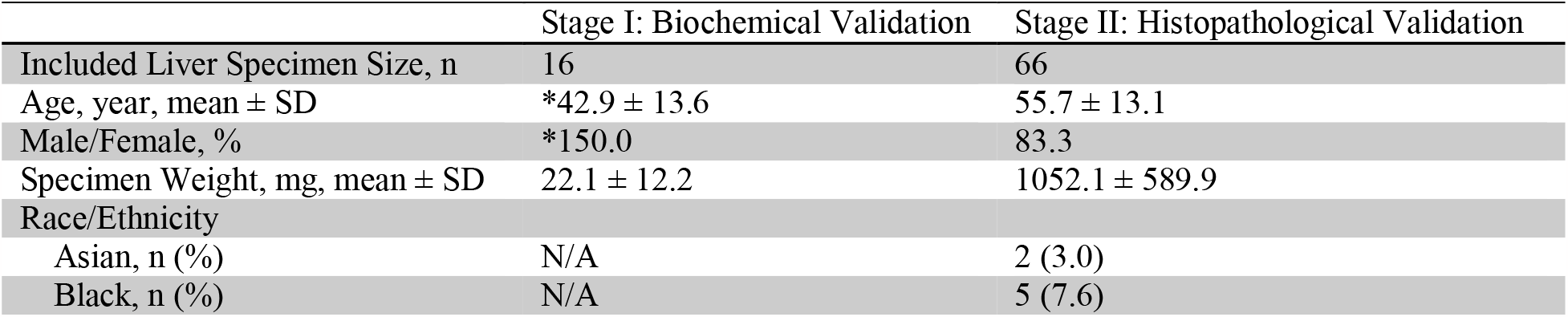
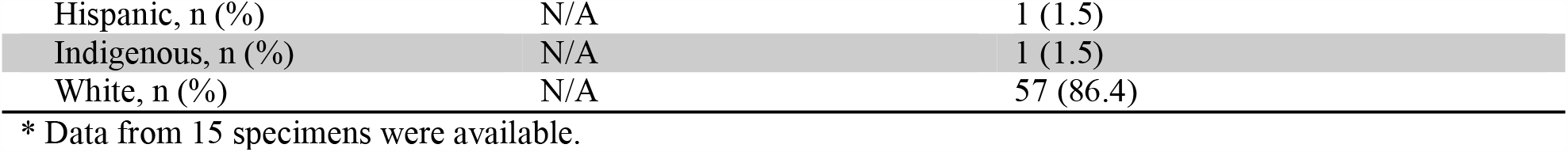
Population characteristics of included human liver specimens.

**Figure 3** shows that the fat contents obtained from the Raman system measurements are positively correlated with triglyceride levels. These correlations imply that signals of both the reflectance and Raman channels contained information about fat contents in human specimens. It should be noted that the reflectance channel signals had a worse linear correlation coefficient than the Raman channel (0.64 vs. 0.82). Typical liver specimens with minimum triglyceride contents between 0.2 μg/μg protein and 0.4 μg/μg protein were predicted to have more than 40% fat in content, implying that the calibration using duck fat-agar phantoms resulted in a significant zero-point error of the reflectance channel.

**Figure 3.**
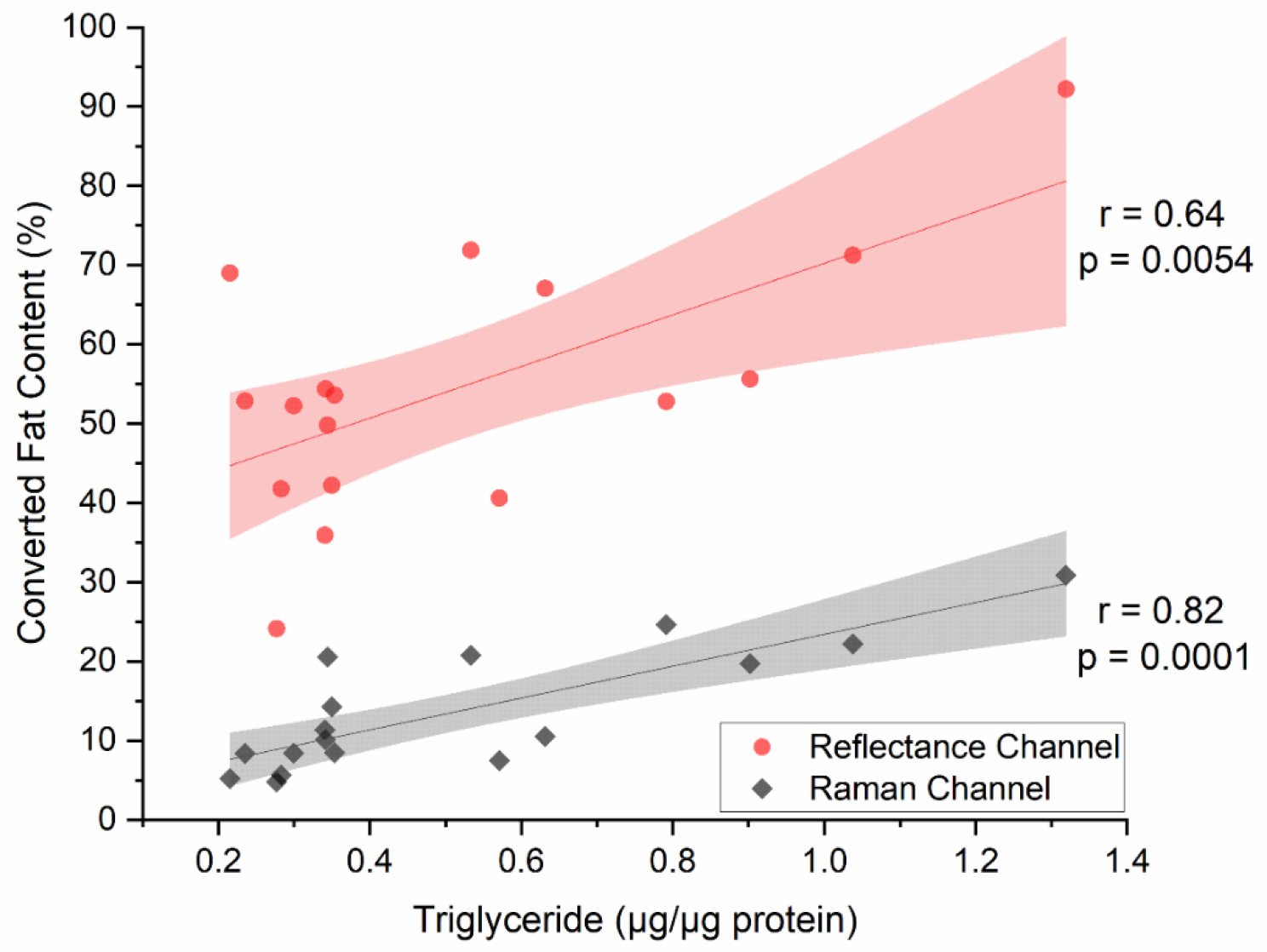
Plots and linear regressions of triglyceride content vs. converted fat contents from voltage signals of the reflectance and Raman channels. Shadow areas: 95% confidence bands of linear regressions.

An expert liver pathologist (AES) assessed the whole slide images (as shown in **Figure 4A**) and estimated, to the nearest 5% interval, the percent of large droplet MaS and the percent of global (macro- and micro-vesicular) HS. The assessment of large droplet MaS was performed per the most recent Banff recommendations^12^ for “large droplet fat,” defined as a single droplet expanding the hepatocyte and larger than adjacent cells, with low power determination of the percentage of tissue affected, followed by higher magnification assessment of fatty areas to determine what proportion of steatotic cells meet the definition of “large droplet fat,” and finally adjustment of the low power percentage accordingly. After completion of pathologist estimates, positive pixels (as shown in **Figure 4B**) were counted automatically using the Positive Pixel Count algorithm v9 of Aperio ImageScope (Leica Biosystems); this value was normalized by the area assessed to generate a percentage. Areas of artifact, such as tissue folding or coverslip lifting, were manually excluded from the analysis.

**Figure 4.**
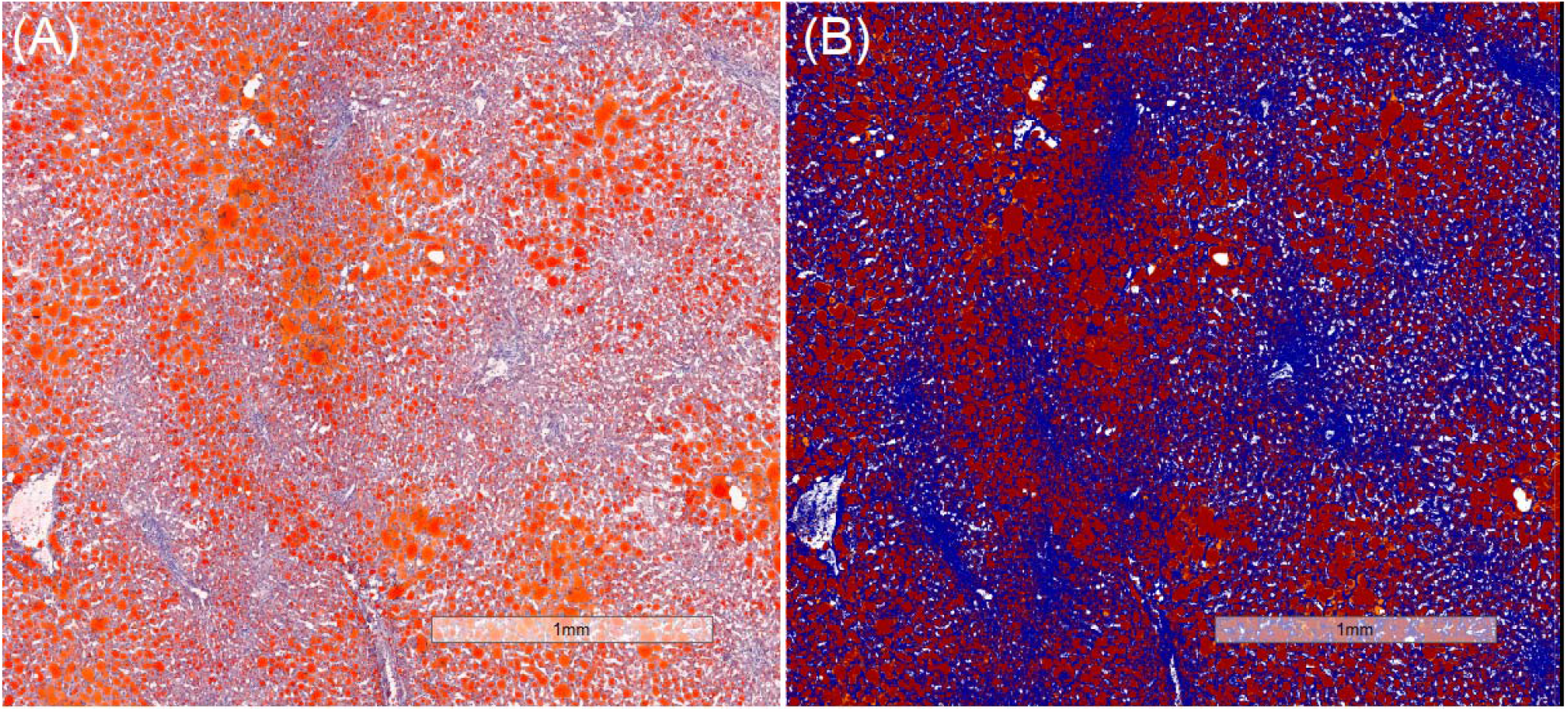
(A) Original view and (B) view after applying the Positive Pixel Count algorithm v9 of digitized images of an ORO-stained frozen section at 40X magnification. The expert pathologist (AES) assessed global HS and MaS based on the original view, and the AI-assessed AI algorithm quantified the ORO stain percentage based on the stained slide pixel area. The ORO-stained lipid droplets (positive pixels) are in orange (A) in brightfield and in red (B) after the application of the Positive Pixel Count algorithm v9.

Considering the better linear correlation of the Raman channel signal with the TG results in the biochemical validation stage, we adopted the Raman channel signal as the primary variable for predicting HS in the histopathological validation stage. The boxplots in **Figure 5** show the distribution of fat content estimated by the Raman system, grouped by the degree of large droplet MaS (assessed by the expert pathologist), ORO stain percentage (estimated by AI), and total steatosis percentage (evaluated by the expert pathologist).

**Figure 3.**
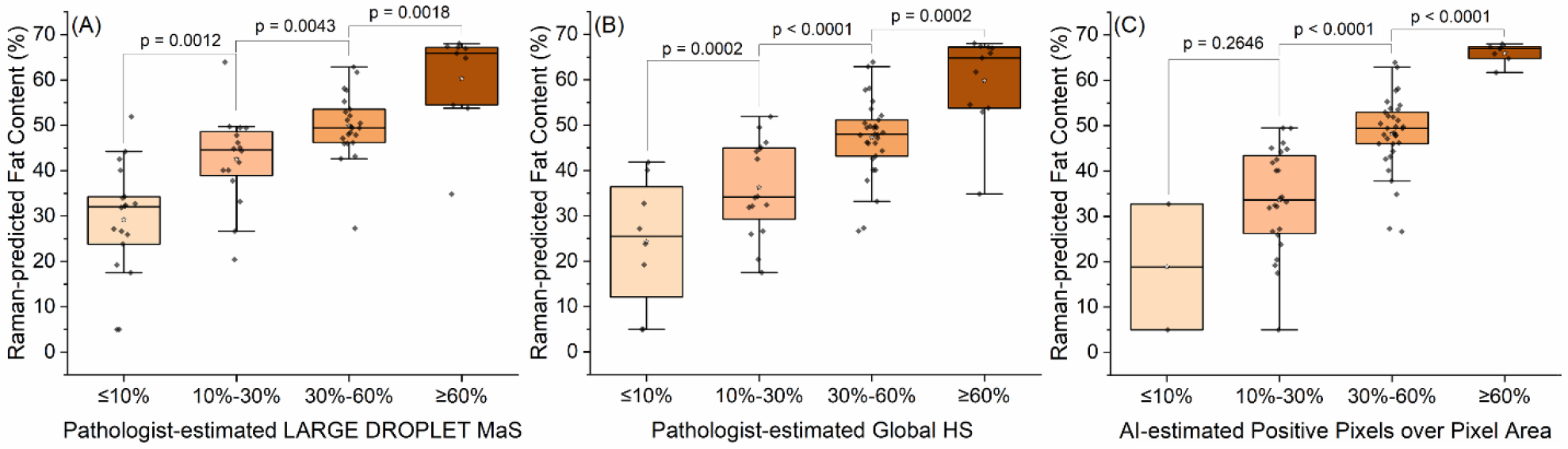
Boxplots of Raman-estimated fat content vs. (A) grade of pathologist-estimated large droplet macrosteatosis, (B) pathologist-estimated all steatosis in percentage (to the nearest 5%), and (C) AI-estimated Oil Red O-stained pixels over pixel area (of Oil Red O-stained slides) in percentage. The boxes represent the interquartile range, with the horizontal lines inside the box indicating the median fat content values. The blank stars inside the box represent the mean fat content values. The whiskers extend from the boxes to represent the minimum and maximum fat content values, excluding any outliers (the 1.5xIQR rule applied) plotted as individual dots outside the whiskers.

Logistic regression (using the “**logit**” command) revealed significant (all p-values < 0.0001) differences in the Raman-estimated fat content among all groups. As shown in **Table 2**, the Raman-estimated fat content, as a single predictor (using the “**predict**” command), effectively differentiated minimum risk (≤10%), low-risk (10%-30%), high-risk (30%-60%), and maximum-risk (≥60%) MaS and global HS, with areas of the operating characteristic curve (AUROC) between 0.88 and 0.90.

**Table 2.** AUROCs of the binary predictions of the Raman channel in differentiating between specimens of MaS, (pathologist-estimated) global HS, and (AI-estimated) ORO stain percentage based on stained slide pixel area at different thresholds. CI: confidence interval.

| Threshold | MaS | 95% CI | Global HS | 95% CI | Positive Pixel Area | 95% CI |
| --- | --- | --- | --- | --- | --- | --- |
| 10% | 0.90 | [0.82-0.99] | 0.90 | [0.80-0.99] | 0.89 | [0.70-1.00] |
| 30% | 0.88 | [0.80-0.96] | 0.88 | [0.80-0.96] | 0.88 | [0.80-0.96] |
| 60% | 0.90 | [0.75-1.00] | 0.90 | [0.75-1.00] | 0.90 | [0.75-1.00] |

Utilizing both Raman and reflectance signals enhanced the AUROC of almost all binary classifications; however, the differences between the Raman and dual-channel predictions were insignificant (see **Table S1**).

Most specimens in the pathological validation stage had a mixed form of MaS and MiS. As Figure **6(A-B)** shows, the degree of MaS of the studied liver specimens was strongly positively correlated with the degrees of pathologist-estimated global HS and AI-estimated positive pixel area, limiting examinations of the investigated Raman system on distinguishing MaS from global HS.

**Figure 4.**
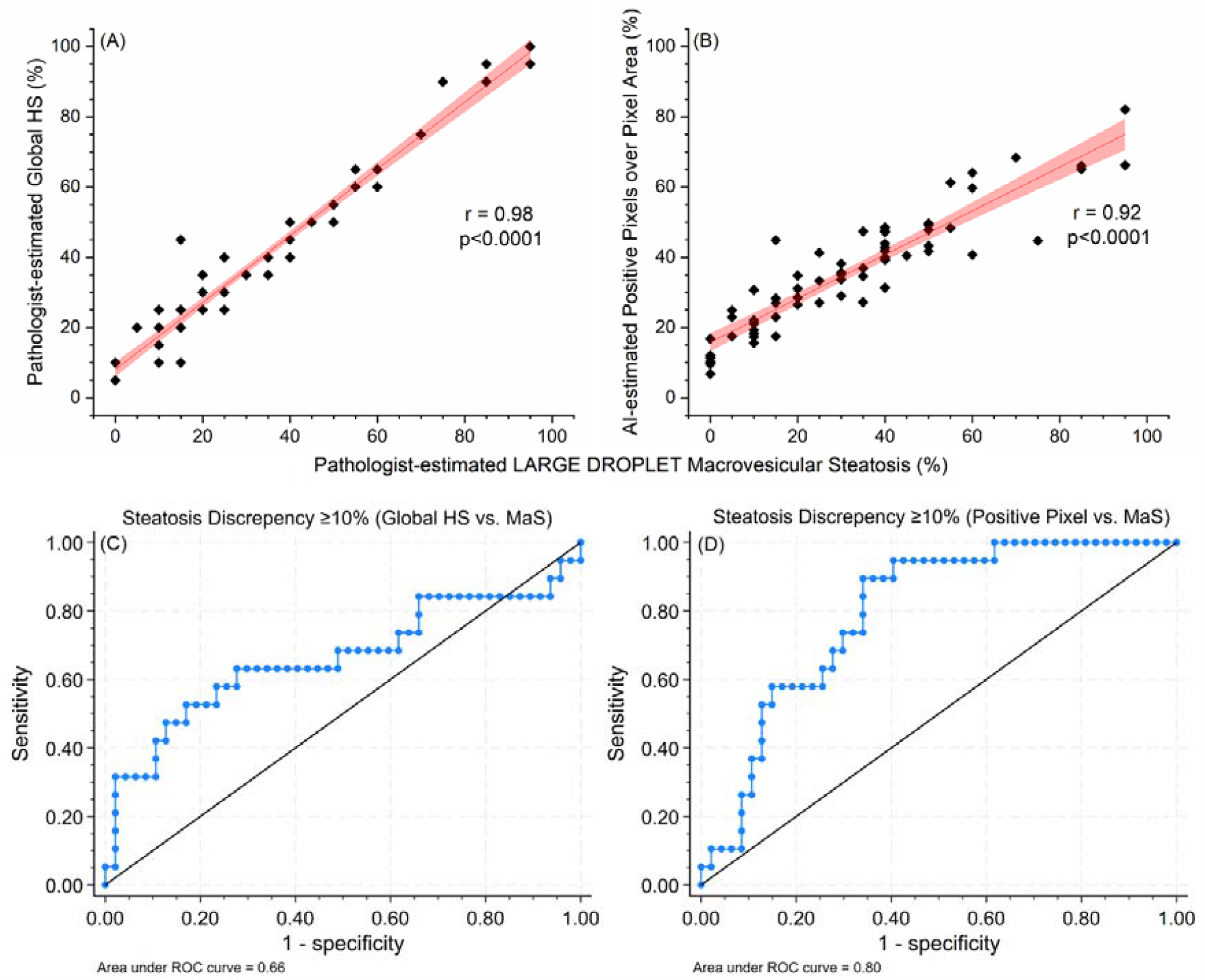
(A-B) Plots and linear regressions of (A) pathologist-estimated global HS vs. pathologist-estimated MaS and (B) AI-estimated global HS vs. pathologist-estimated MaS. (C-D) ROCs of dual-variable binary predictions on ≥10% steatosis discrepancy between (C) pathologist-estimated global HS and pathologist-estimated MaS and (D) AI-estimated global HS and pathologist-estimated MaS. The Raman and the reflectance channels were applied as dual predictors.

Considering the inherently high correlation with global HS (assessed by the expert pathologist and AI algorithm) and the pathological assessments are to the nearest 5% steatosis, we set a compromised steatosis discrepancy threshold at 10%. Information from the reflectance channel was paired with the Raman-estimated fat content for dual-variable predictions. **Figure 6(C)** shows the ROC curves of binary predictions on ≥10% steatosis discrepancy between pathologist-estimated global HS and pathologist-estimated MaS, with an AUROC of 0.66. This could be due to data points’ degradation (overlapping), as the expert pathologist’s estimations were to the nearest 5%.

The ROC curve of binary predictions on ≥10% steatosis discrepancy between AI-estimated global HS (positive pixel area) and pathologist-estimated MaS presented a better AUROC of 0.80, as shown in **Figure 6(D)**. This may be attributed to the AI-based estimation of global HS, which quantified the areas of ORO-stained lipid droplets in liver specimens, yielding more precise percentage readings.

## 4 Discussion

The accumulation of TGs within hepatocytes is characteristic of HS.^35^ Raman spectroscopy has been validated to assess liver steatosis in animal models based on characteristic Raman peaks.^20,21^ While the calibration of the investigated Raman system was in keeping with previous reports,^26^ the investigated Raman system demonstrated substantial positive correlations with both Raman (r = 0.82) and reflectance (r = 0.64) channels, indicating its capability to assess TG in liver specimens. This made it suitable for further evaluation in the histopathological validation stage.

In the histopathological validation stage, the Raman channel was revealed to be able to classify MaS and global HS of human liver specimens. Combining information from the two channels, dual-variable predictions of 66 liver specimens reported good (AUROC = 0.80) classification of an ≥10% steatosis discrepancy between pathologist-estimated MaS and AI-estimated HS (ORO-stained pixel area). These findings indicated that the investigated ambient light-compatible Raman system was capable of accurate ex-situ evaluation of global HS in human livers. The investigated Raman system has demonstrated preliminary efficacy in MaS assessments, indicating its potential as a valuable tool for the in-situ real-time surgical evaluation of donor liver steatosis.

The primary strength of the present study lies in the multifaceted correlation of Raman assessments with a robust array of subjective and objective references. This included TG quantification, expert pathologist evaluation, and advanced AI-based estimation, achieving validations from diverse and complementary perspectives. Moreover, the observed steatosis within the specimens in the histopathological validation stage exhibited a broad distribution of minimum-to-severe steatosis, facilitating a comprehensive evaluation of the HS classifications through the investigated Raman system. This wide-ranging analysis underscores the potential of our approach in advancing the understanding of integrated spectroscopic evaluation on HS and may pave the way for surgical diagnostic techniques.

All specimens included in this study were initially snap-frozen. Although the specimens could not represent donor livers for recovery, their chemical compositions (to which Raman spectroscopy is sensitive) and morphology were to a great extent comparable to those of donor liver organs. All the specimens studied in the pathological validation stage were several times thicker than the 1-mm laser penetration depth reported in the previous clinical trial,^26^ enabling enough scattering interaction as a shone liver organ would provide.

This study encountered several limitations in evaluating the Raman system for HS assessment. A significant hurdle was the mixed presence of MaS and MiS in most specimens, complicating the distinction between MaS and global HS. During the biochemical validation stage, the exclusion of 9 out of 25 specimens due to their size and weight markedly diminished the statistical power, and the reliance on duck fat-agar phantoms for setting up a “zero point” by calibration hampered the system’s accuracy and convenience.

The research noted a promising dual-variable prediction based on Raman scattering and reflectance, capable of classifying specimens with a minimum 10% greater global HS than MaS. However, this finding, lacking a solid physicochemical basis, necessitates further exploration. van Staveren et al. (1991) accurately predicted the scattering coefficient of fat emulsion lntralipid-10% using Mie theory and the size distribution of lipid particles.^36^ A reasonable guess is that the reflectance channel (Mie scattering) contained information about both fat content and lipid droplet size; however, more studies according to the physical principles of Raman and Mie scattering, e.g., Monte Carlo simulations, are needed to probe the concept of the dual-variable prediction. Additionally, the untested performance of the Raman system in operating room conditions, despite its proven resistance to 10,000 lm LED light in laboratory settings, leaves a significant gap in understanding its comprehensive compatibility.^26^

In this study, a single expert pathologist (AES) assessed all microscopic slide images and estimated the percentage of large droplet MaS and global HS. Despite the potential for diverse insights from multiple pathologists,^37,38^ we opted for a single expert to ensure consistent assessment methods and mitigate the risk of interobserver discordance. This approach also assured the quality of the assessment, leveraging the pathologist’s high expertise. The Positive Pixel Count algorithm, as an additional independent approach to quantify the fat content, validated the pathologist’s estimations and enabled finer quantitive discrepancy of global HS and MaS.

To the best of the authors’ knowledge, the present study represents the inaugural trial of conventional Raman spectroscopy on human liver specimens for HS assessment. Advancing beyond the scope of preceding Raman studies, this research emphasized an ambient light-compatible methodology. The efficacy of the Raman spectroscopic approach in assessing the HS of human liver specimens was validated by correlating the fat content results provided by the examined Raman system with TG quantifications. Additionally, histopathological validation was conducted across a wide range of steatotic human liver specimens, culminating in the development of a dual-variable prediction for significant discrepancies (>10%) between global HS and MaS. The insights gained from this study may contribute to enhancing other spectroscopic surgical instruments.

## 5 Conclusion

This research marks a novel endeavour in utilizing a portable spectroscopic system to assess hepatic steatosis in human liver specimens. Two-stage biochemical and histopathological validations highlight the potential of the examined system as a trustworthy, non-invasive modality for steatosis evaluation.

Notably, the ambient light-compatible approach signifies a notable progression beyond prior spectroscopic methods, effectively addressing associated limitations. Raman scattering was validated able to quantify lipid content and therefore detect the presence of Global HS in human liver specimens.

Moreover, introducing a dual-variable prediction for identifying significant (≥10%) discrepancies in global HS and MaS demonstrates the potential of differentiating between steatotic livers with different extent of MaS.

Beyond immediate outcomes, this study lays the foundation for the broader application of ambient light-compatible spectroscopic probes in clinical environments, potentially revolutionizing intraoperative liver assessments. Amidst the escalating demand for liver transplantation, tools of this nature hold substantial promise in ensuring optimal graft quality, consequently benefiting liver recipients. While the results are encouraging, further research is imperative to affirm the comprehensive potential and applicability of this system, especially in real-world operating room contexts.

## Data Availability

All data produced in the present study are available upon reasonable request to the authors

## Acknowledgement

The authors thank Menno Hoekstra for training H.G. and Y.S.C. for triglyceride quantification of liver tissues. The authors also thank Lab2Market, Ready2Launch, and ideaHUB for vital technical and financial support in developing and iterating our prototype systems.

## Abbreviations used in this paper

AI: artificial intelligence
AUROC: area under the operating characteristic curve
HS: hepatic steatosis
LT: liver transplantation
LUMC: Leiden University Medical Center
MaS: macrovesicular steatosis
MiS: microvesicular steatosis
OR: operating room
ORO: Oil Red O
ROC: receiver operating characteristic
SD: standard deviation
TG: triglyceride
VUMC: Vanderbilt University Medical Center

## Ethics Statement

Ethical review and approval were not required for the study at Leiden University Medical Centre on human liver specimens by the local legislation and institutional requirements. The Institutional Review Board at Vanderbilt University approved the study at Vanderbilt University (IRB #: 121423). Both studies were approved by the Health Sciences Research Ethics Board at Dalhousie University (REB #: 2022-6133 and 2022-6318).

## Declaration of AI and AI-assisted technologies in the writing process

Statement: During the preparation of this work, the authors used ChatGPT-4 (Open AI, San Francisco, CA, USA) in order to improve readability and language. After using this tool, the authors reviewed and edited the content as needed and take full responsibility for the content of the publication.

## Supplementary materials

**Figure S5.**
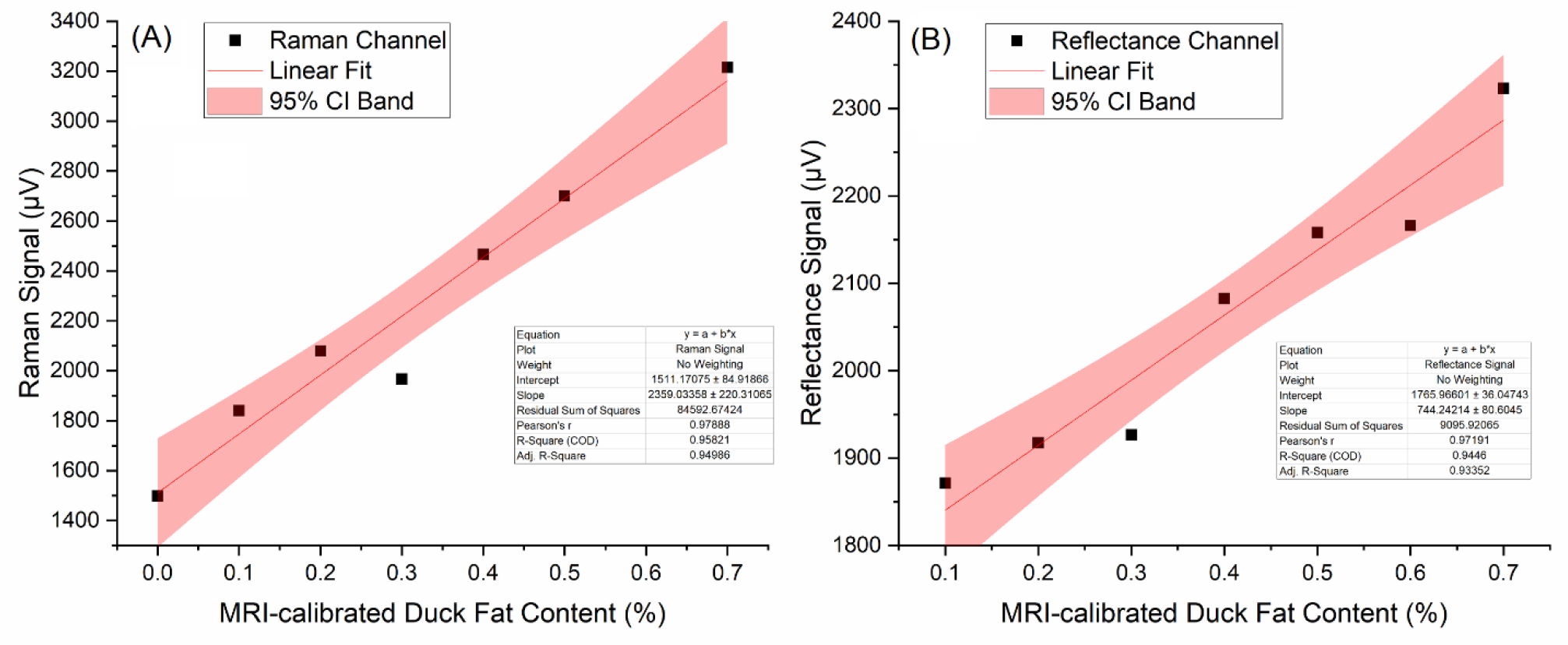
Examples of calibrations using the duck fat phantoms. (A) The Raman channel; (B) the reflectance channel

**Table S1.** Comparisons among single- and double-variable classification methods. *p-value < 0.05, indicating the compared AUROCs are statistically significantly different.

| Threshold | Area Under ROC Curve | | | p-value ($H_0$: Equal AUROC) | | | |
| --- | --- | --- | --- | --- | --- | --- | --- |
|  | Raman (#1) | Reflectance (#2) | Dual (#3) | #1, 2 | #1, 3 | #2, 3 | #1, 2, 3 |
| **Macrosteatosis** |  |  |  |  |  |  |  |
| 10% | 0.90 [0.82-0.99] | 0.84 [0.91-0.96] | 0.94 [0.88-0.99] | 0.381 | 0.204 | 0.066 | <0.001* |
| 30% | 0.88 [0.80-0.96] | 0.84 [0.75-0.94] | 0.91 [0.84-0.98] | 0.564 | 0.153 | 0.085 | 0.003* |
| 60% | 0.90 [0.75-1.00] | 0.75 [0.54-0.95] | 0.89 [0.73-1.00] | 0.151 | 0.532 | 0.151 | 0.336 |
| **All steatosis** |  |  |  |  |  |  |  |
| 10% | 0.90 [0.80-0.99] | 0.78 [0.57-1.00] | 0.91 [0.82-1.00] | 0.365 | 0.552 | 0.277 | 0.214 |
| 30% | 0.88 [0.80-0.96] | 0.83 [0.71-0.94] | 0.91 [0.85-0.98] | 0.491 | 0.260 | 0.104 | 0.016* |
| 60% | 0.91 [0.79-1.00] | 0.79 [0.61-0.96] | 0.90 [0.78-1.00] | 0.170 | 0.702 | 0.139 | 0.196 |
| **Stained pixels** |  |  |  |  |  |  |  |
| 10% | 0.89 [0.70-1.00] | 0.97 [0.92-1.00] | 0.98 [0.94-1.00] | 0.388 | 0.378 | 0.480 | 0.670 |
| 30% | 0.90 [0.83-0.97] | 0.79 [0.67-0.91] | 0.91 [0.84-0.98] | 0.093 | 0.657 | 0.025 | 0.021* |
| 60% | 0.99 [0.98-1.00] | 0.88 [0.75-1.00] | 1.00 [1.00-1.00] | 0.091 | 0.413 | 0.070 | 0.084 |

